# Detection and isolation of asymptomatic individuals can make the difference in COVID-19 epidemic management

**DOI:** 10.1101/2020.04.23.20077255

**Authors:** Lía Mayorga, Clara García Samartino, Gabriel Flores, Sofía Masuelli, María V. Sánchez, Luis S. Mayorga, Cristián G. Sánchez

**Affiliations:** Instituto de Histología y Embriología de Mendoza (IHEM, Universidad Nacional de Cuyo, CONICET)-Centro Universitario UNCuyo, (5500) Mendoza, Argentina; Instituto de Química Biológica de la Facultad de Ciencias Exactas y Naturales (IQUIBICEN) CONICET, Ciudad Universitaria, C1428EGA, Ciudad Autónoma de Buenos Aires, Argentina; Facultad de Ciencias Médicas, Universidad Nacional de Cuyo-Centro Universitario UNCuyo, 5500 Mendoza, Argentina; Eventbrite Company (5500) Mendoza, Argentina; Instituto de Medicina y Biología Experimental de Cuyo (IMBECU), Centro Científico y Tecnológico de Mendoza (CCT-Mendoza), CONICET, UNCuyo, (5500) Mendoza, Argentina; Facultad de Ciencias Exactas y Naturales, Universidad Nacional de Cuyo, (5500) Mendoza, Argentina; Instituto Interdisciplinario de Ciencias Básicas, Universidad Nacional de Cuyo, CONICET, Facultad de Ciencias Exactas y Naturales, Padre J. Contreras 1300, (5500) Mendoza, Argentina

**Keywords:** COVID-19, SARS-Cov-2, SEIR mathematical modelling, Asymptomatic, healthcare burden

## Abstract

Mathematical modeling of infectious diseases is a powerful tool for the design of management policies and a fundamental part of the arsenal currently deployed to deal with the COVID-19 pandemic. Here we present a compartmental model for the disease where symptomatic and asymptomatic individuals move separately. We introduced healthcare burden parameters allowing to infer possible containment and suppression strategies. In addition, the model was scaled up to describe different interconnected areas, giving the possibility to trigger regionalized measures. It was specially adjusted to Mendoza-Argentina’s parameters, but is easily adaptable for elsewhere. Overall, the simulations we carried out were notably more effective when mitigation measures were not relaxed in between the suppressive actions. Since asymptomatics or very mildly affected patients are the vast majority, we studied the impact of detecting and isolating them. The removal of asymptomatics from the infectious pool remarkably lowered the effective reproduction number, healthcare burden and overall fatality. Furthermore, different suppression triggers regarding ICU occupancy were attempted. The best scenario was found to be the combination of ICU occupancy triggers (on: 50%, off: 30%) with the detection and isolation of asymptomatic individuals. In the ideal assumption that 45% of the asymptomatics could be detected and isolated, there would be *no* need for quarantine, and Mendoza’s healthcare system would not collapse. Our model and its analysis inform that the detection and isolation of *all* infected individuals, without leaving aside the asymptomatic group is the key to surpass this pandemic.

## INTRODUCTION

The COVID-19 pandemic has brought the world to a pause with the sole aim to defeat this worldwide threat, and the scientific community has joined the effort. Since its outbreak by the end of 2019, we have been able to learn some about this new SARS-Cov2 coronavirus. Unfortunately, new facts come with a lag compared to the virus spread and governments are forced to make prompt decisions based on limited evidence which changes at a staggering pace. The fight against a practically unknown enemy has been and still is the major obstacle. Aside from studying the virus’s biology, infecting mechanisms, probable treatments and of course vaccine development, epidemic mathematical modelling has stepped forward.

In a trade-off between simplicity and detail, compartmental modelling strategies provide a sharp suit that allows exploring a variety of scenarios and provides an intuitive understanding of the most critical factors governing disease dynamics. The recent use of S(susceptible)-Exposed(E)-Infected(I)-Recovered(R) models has made a difference for public health care decision making by providing, for example, estimations of the impact of Non-Pharmaceutical Interventions (NPI)[1,2]. The main challenge is to create a model that predicts plausible scenarios for a disease we have known for only four months.

One of the most important barriers for the provision of solid epidemiological parameters has been the different management strategies that each country has taken in response to this outbreak. Most evidently, the Case fatality Rate (CFR) varies largely between countries (i.e. Italy 12%, Argentina 3%, Iceland 0.3%). Varying CFRs cannot be explained only by the different population age structure or available critical care beds. Most importantly, uneven and time-varying testing criteria in different countries make the CFR an inadequate severity parameter. South Korea and especially and more recently, Iceland’s approach to testing massively for COVID-19 has brought us closer to the real Infection Fatality Rate (IFR) which describes more precisely the magnitude of the threat [3-5]. Better estimates of the IFR have given insights on two aspects. Firstly, the asymptomatic or very mildly symptomatic group of individuals is more significant than previously thought[6,7] since they represent the vast majority of the infected individuals[4,5,8]. Secondly, these individuals, in most cases, are not detected nor isolated, and therefore appear to be the leading cause of the epidemic’s spread. We proposed ourselves to model the strike of the virus locally (Mendoza-Argentina). Anyhow, our model applies to any city or country and available for use and adaptation. Argentina, as a developing country, was not going to be able to bear this pandemic without a health care collapse. Based on the epidemic behaviour in Europe, the government determined a complete lockdown as the primary measure of control for the country as from March 20th, when Argentina’s confirmed positive cases were only 128, mainly located in the capital city Buenos Aires and mostly imported. Many regions from inside Argentina had zero confirmed cases, including the authors’ hometown, Mendoza province. Prompt lockdown measures significantly flattened the curve at an early stage. Arguably this was an anticipated measure, and it gave time to prepare (at least to some extent) for what was/is coming.

As mentioned by Ferguson et al.[9], the efficiency of mitigation and suppression measures depends on the size of the country or region in which these actions are implemented. Different population densities, uneven access to intensive health care, distinguishing age-related communities, all make the epidemic spread distinctively. Therefore, a model should be able to take these variables into account.

We propose here an SEIR model for COVID-19 epidemics that incorporates specific compartments that classify infected individuals in several clinical categories. These compartments provide figures that can inform the strategic planning of health care requirements. On the other hand, different regions exchange infectious and exposed individuals through communication routes. Consequently, the model can provide the possibility to trigger measures independently for each city and block intercommunications selectively. Most importantly, we model asymptomatic individuals as a subset of the infectious compartment. We demonstrate here the significant impact that detecting and isolating these individuals can have on the disease outcome.

## METHODS

We augmented the basic SEIR scheme by modelling symptomatic and asymptomatic individuals separately. Symptomatic individuals can move into mild and severe cases which can recover or further evolve into critical care and recover or die. Asymptomatic individuals may move into an isolated compartment after a detection lag of variable efficiency. Fig. 1 shows a connection diagram of the compartments in our model and Table 1 provides a detailed description of each. Fig. 2 shows the possible timelines of the evolution of an initially susceptible individual together with the relevant parameters that determine the flow between compartments. Table 1 provides a detailed description of each compartment.

**Fig.1:**
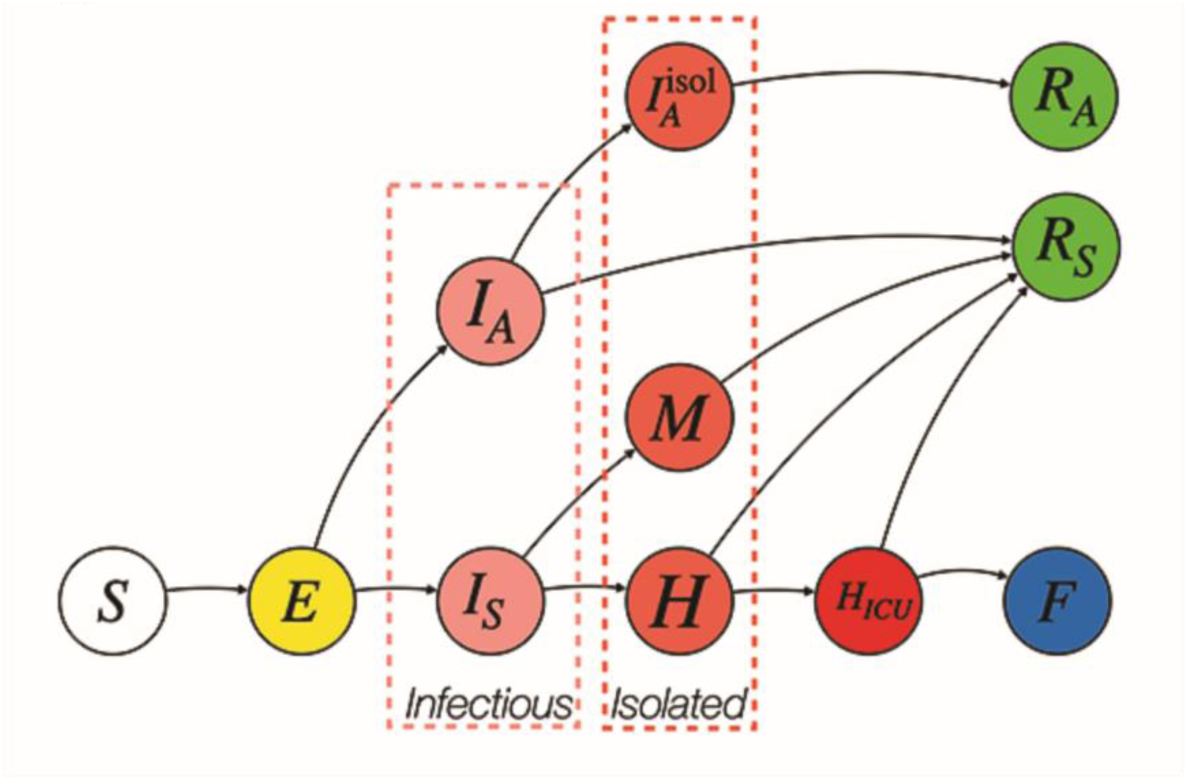
Compartments and connections in the modified SEIR model we propose.

**Table 1:** The model’s compartments

| Compartment | Meaning |
| --- | --- |
| $S$ | Susceptible |
| $E$ | Exposed |
| $I_A$ | Infected asymptomatic |
| $I_S$ | Infected symptomatic |
| $I_A^{isol}$ | Isolated asymptomatic |
| $R_A$ | Recovered from asymptomatic cases |
| $M$ | Mild |
| $H$ | Hospitalized |
| $H_{ICU}$ | In critical care |
| $R_S$ | Recovered from symptomatic cases |
| $F$ | Deceased |

**Fig.2:**
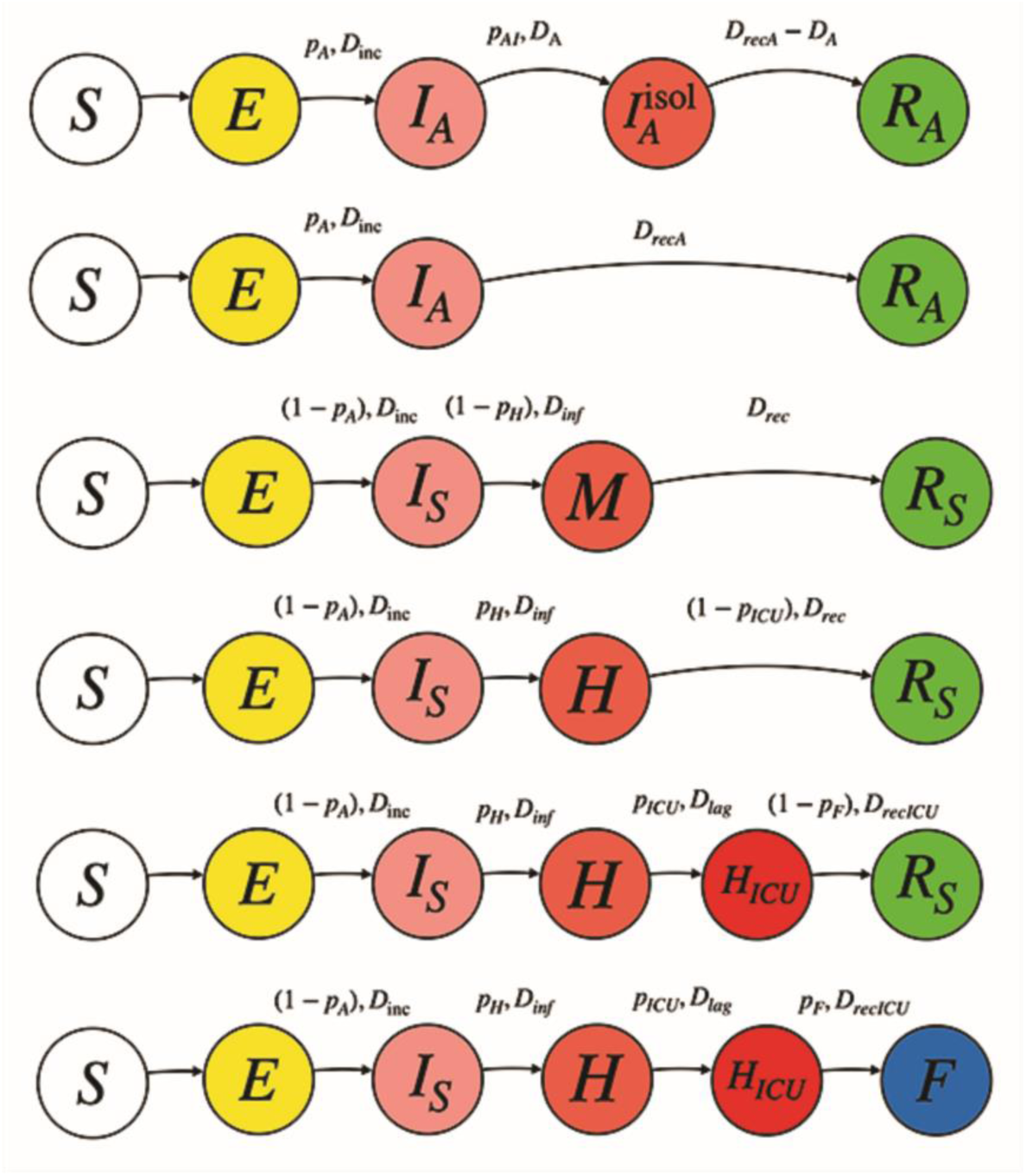
Possible timelines for an initially susceptible individual. Over the arrows connecting each compartment we show the relevant mean residence times and branching probabilities. The time and probabilities we used for the analysis are stated according to the literature in Table 2.

Based on recent calculations on the IFR, of 0.39-1.33% [3], and most recently of 0.01-0.19% inferred from Iceland’s statistics[4,5], we adapted the parameters for our model to our country. Taking in consideration an IFR of ~0.3% and the current CFR in Argentina of ~3% we concluded that 90% of the infected cases were asymptomatic or with very mild symptoms not fulfilling the current Argentine criteria for COVID-19 testing (fever & sore throat or cough or respiratory distress). The asymptomatic case percentage is in line with the published data by Li et al. [8] who estimated the undocumented cases in 86%.

Argentina’s principle to test only highly suspicious cases is leaving out asymptomatic and paucisymptomatic individuals. For our model, we used the complete epidemiological data we found[3,9] that referred to a subset of patients from China for which a similar criterion for COVID-19 testing and hospitalization than Argentina was used. We adapted it to maintain the CFR of ~3% and IFR~0.3%. Summarizing, from what is called “symptomatic cases” in Argentina today, around 10% will require hospital care, of those, 36% will need intensive care unit facilities, and of the latter, 50% will die.

Based on European case growth rates, and in agreement with the parameters we set for our model, we presumed a basic reproduction number of 4[2]. We assumed that mitigation measures (case-isolation, general social distancing, banning of public gatherings, university closures) lower *R_0_* to 2 and suppression measures (complete lockdown or quarantine of the whole population except for essential activities) could make *R*_0_≤1[2]. We supposed, as published, that asymptomatic individuals are half as infective as symptomatic patients[8,9]. We inferred for these an attenuation factor of 0.5 in the basic reproduction number and ⅓-½ the infective time of symptomatic cases[9,10]. Table 2 shows the parameters we chose and the bibliography that supports our choices.

**Table 2.** The model’s parameters

Parameters used for our SEIR model, their value and the bibliography that supports them.
| Parameter | Meaning | value | References |
| --- | --- | --- | --- |
| $R_0$ | Basic reproduction number | 4 | Flaxman et al. .2020 [2] |
| $D_{inc}$ | Incubation period (days) | 5 | Flaxman et al. .2020 [2] |
| $D_{inf}$ | Time for which a symptomatic individual is infectious (days) | 4 | Li et al. 2020[8] |
| $D_{rec}$ | Time to recover for $M$ and $H$ cases (negativize PCR) that do not require ICU (days) | 14 | Fang et al. 2020[11] |
| $D_{recA}$ | Time to recover (negativize PCR) for an $I_A$(days) | 7 | Zhang et al. 2020[10,12] |
| $D_{lag}$ | Time for $H$ to require ICU (days) | 7 | [6] |
| $D_{recICU}$ | Time for $H_{ICU}$ to recover in ICU (days) | 14 | Fang et al. 2020[11] |
| $p_A$ | Probability of an infected patient to be asymptomatic | 0.9 | Li et al. 2020[8] |
| $p_H$ | Probability of a symptomatic individual to require hospitalization | 0.1 | Verity et al. 2020, Ferguson et al. 2020[3,9] |
| $p_{ICU}$ | Probability of $H$ to require ICU | 0.36 | Verity et al. 2020, Ferguson et al. 2020[3,9] |
| $p_F$ | Probability of $H_{ICU}$ to die | 0.5 | Ferguson et al. 2020[9] |
| $f_A$ | Attenuation factor for asymptomatic patients | 0.5 | Li et al. 2020[8] |

The SEIR model depicted in Fig. 1 gives rise to the following set of ordinary differential equations:

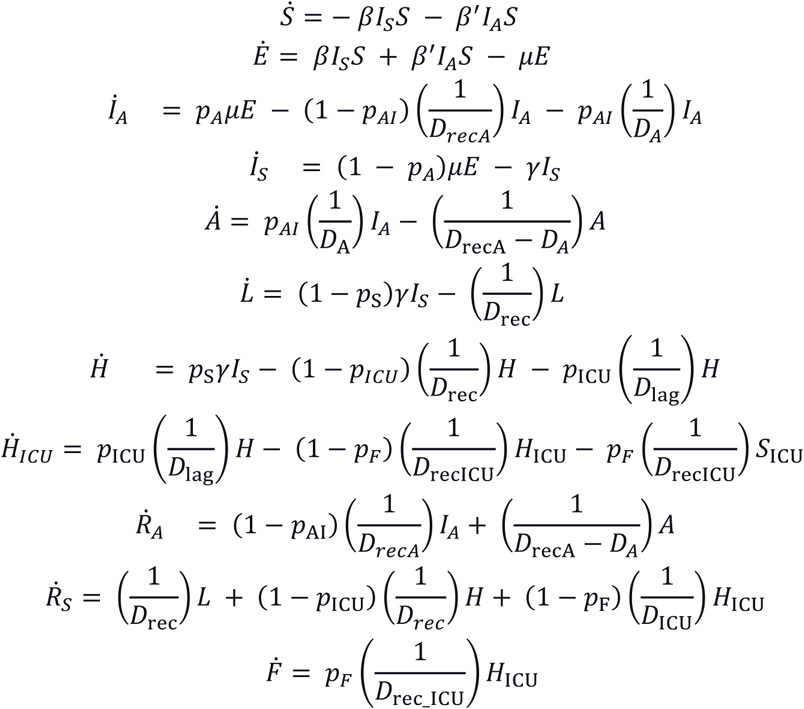

Where dotted quantities are time derivatives and 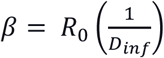, 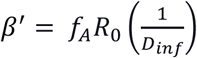, 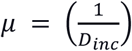 and 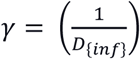. Probabilistic parameters are adjusted to provide a CFR and IFR of 3 and 3% respectively in equilibrium. The quantity *p_AI_* represents the probability that an asymptomatic individual is detected and isolated. The mean residence time parameters govern the dynamical evolution of the compartment populations. The model is scaled up to describe different interconnected areas or cities by integrating the ordinary differential equation set for periods of a day and exchanging exposed and infectious individuals at the end of each day according to the following equation:

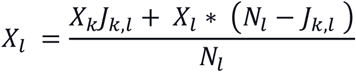

Where *X_l_* and *X_k_* stand for the fraction of exposed and infectious individuals in regions *l* and *k* respectively, *N_l_* is the population of area *l* and *J_kl_* is the number of people exchanged daily between areas *l* and *k*. Infectious individuals include both asymptomatic and symptomatic ones.

We divided the Mendoza province into 4 regions: North (mostly urban), West (vineyard and mountain zone), East (primarily rural) and South (urban and rural). Hospital beds and ICU (intensive care units) for each area were added to the model. Additionally, an estimate of how many people travel daily between the regions was considered (Fig.3).

**Fig.3:**
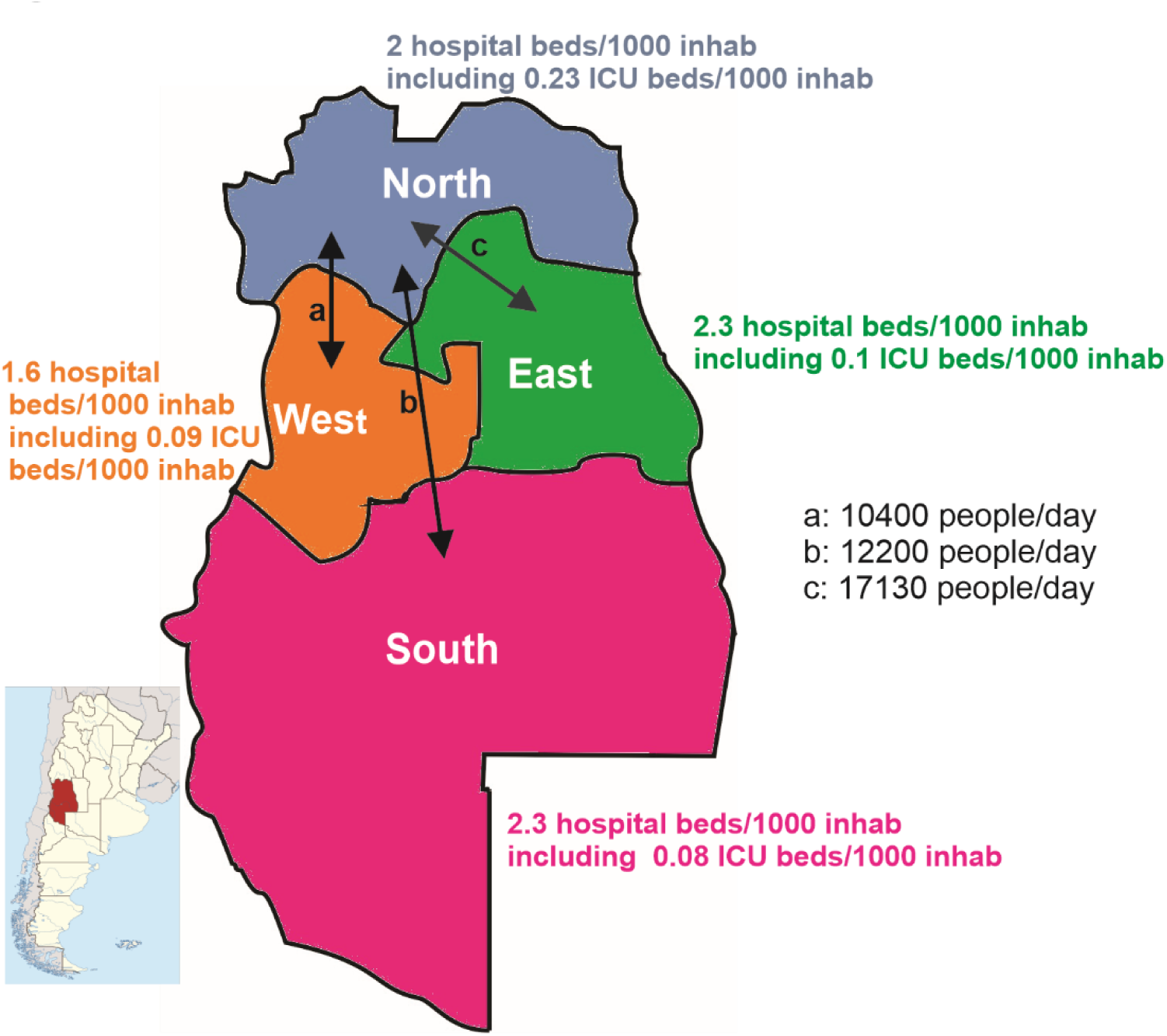
Mendoza province, located in the Center-West of Argentina (shown in red in the Argentina map). We present the four zones in which we compartmentalized the model detailing hospital beds/1000 inhabitants and daily mean people exchange between regions (a, b, c).

## RESULTS

### Use of the regional compartmentalization of the model

We set our model to run in 4 zones of the Mendoza province considering their respective population, daily mobilization of people between the regions, and health care facilities for each area (Fig. 3). As an example of the application of this tool, we simulated an outbreak in the North zone, starting with one infected person and monitored its arrival to the West area. We contemplated two different reproduction numbers; *R*_0_ = 4 (no mitigation nor suppression actions installed) or *R*_0_ *=* 2 (moderate containment measures governing the whole province). Around 10400 people travel every day between these districts. Without interfering in the communication routes between zones, it took 29 days *(R*_0_ = 4) and 88 days *(R*_0_ = 2) for the virus to spread to the West zone. When travelling between the communities was reduced to a tenth, the outbreak reached the West with a 17-day lag (day 46) in the *R*_0_ = 4 scenario and a 42-day delay (day 130) for the *R*_0_ = 2 scenario (Fig.4). As expected, this shows that blocking communication routes between districts is an excellent strategy to delay the entrance of the epidemic. Anyhow, this action has more power when combined with other containment measures as shown here by comparing a *R*_0_ = 4 versus an *R*_0_ = 2 situation. Furthermore, in this restricted communication situation, the exponential curves eventually catch up if no other interventions are instated (data not shown in graph). The communication restriction between North and West practically does not modify the North’s dynamic, so in Fig.4, we only show the basal state of the Northern District.

**Fig. 4:**
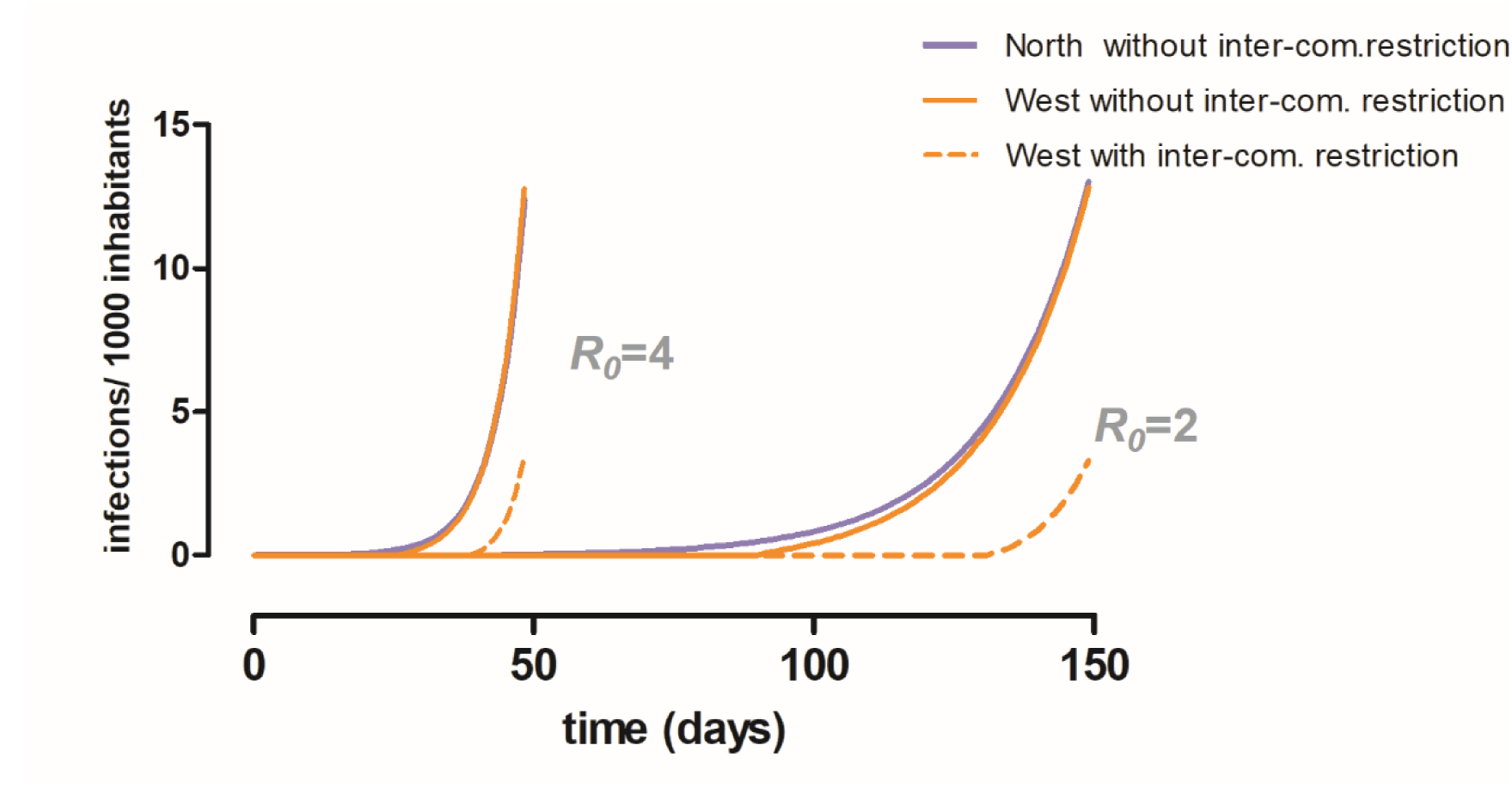
The impact of district intercommunication restriction on the infection spread. We show the infection rate of an outbreak initiated in the Northern District and how it spreads to the West zone with and without a 90% intercommunication restriction policy. Curves for two *R_0_* scenarios are plotted *(R_0_=4* and *R_0_*=2). The communication restriction between North and West practically does not modify the North’s dynamic, so we only show the basal state of the Northern District.

### The importance of the asymptomatic group in the model

Considering that the asymptomatic or very mildly affected individuals are the majority, as can be expected, to detect and isolate them diminishes the total infectious population and changes the epidemic evolution dynamics. As Li et al. have shown[8] the introduction of an asymptomatic reservoir with a different reproduction number modifies the epidemic evolution in a non-trivial manner. This effect comes into place by a modification of the effective reproduction number *(R_e_)* when asymptomatic individuals are considered. Elaborating on this result, we can add that fast and efficient detection plus the isolation of asymptomatic individuals can indeed control the epidemic. The effective reproduction number of the model shown in Fig. 1 is significantly altered by fast and efficient detection plus isolation of asymptomatic individuals, as shown in Fig. 5a. If the efficiency of detection is 50% within three days of becoming infectious, the effective reproduction number can be reduced by a half. If this policy is accompanied by other non-pharmaceutical interventions that lower the basic reproduction number such as mitigation and suppression measures then the effective reproduction number can be brought to values lower than one, effectively controlling the epidemic. On the opposite extreme, the effective reproduction number is larger than the *R_0_* parameter when asymptomatics are not isolated and therefore remain infectious until recovery. For details on the calculation of the *R_e_* please refer to the supplementary online resource material (ESM _1) As can be expected from its influence on the effective reproduction number, the isolation of asymptomatic individuals has a dramatic effect on the duplication time of the epidemic in the exponential growth phase (Fig. 5b). In a basic reproduction number scenario of 2, isolating half of the asymptomatic individuals within four days of becoming infectious can effectively double the time it takes for clinical cases to duplicate in the exponential growth phase. This effect is smaller for more significant reproduction numbers reinforcing the statement that other interventions must accompany this policy in order to control the epidemic.

**Fig. 5:**
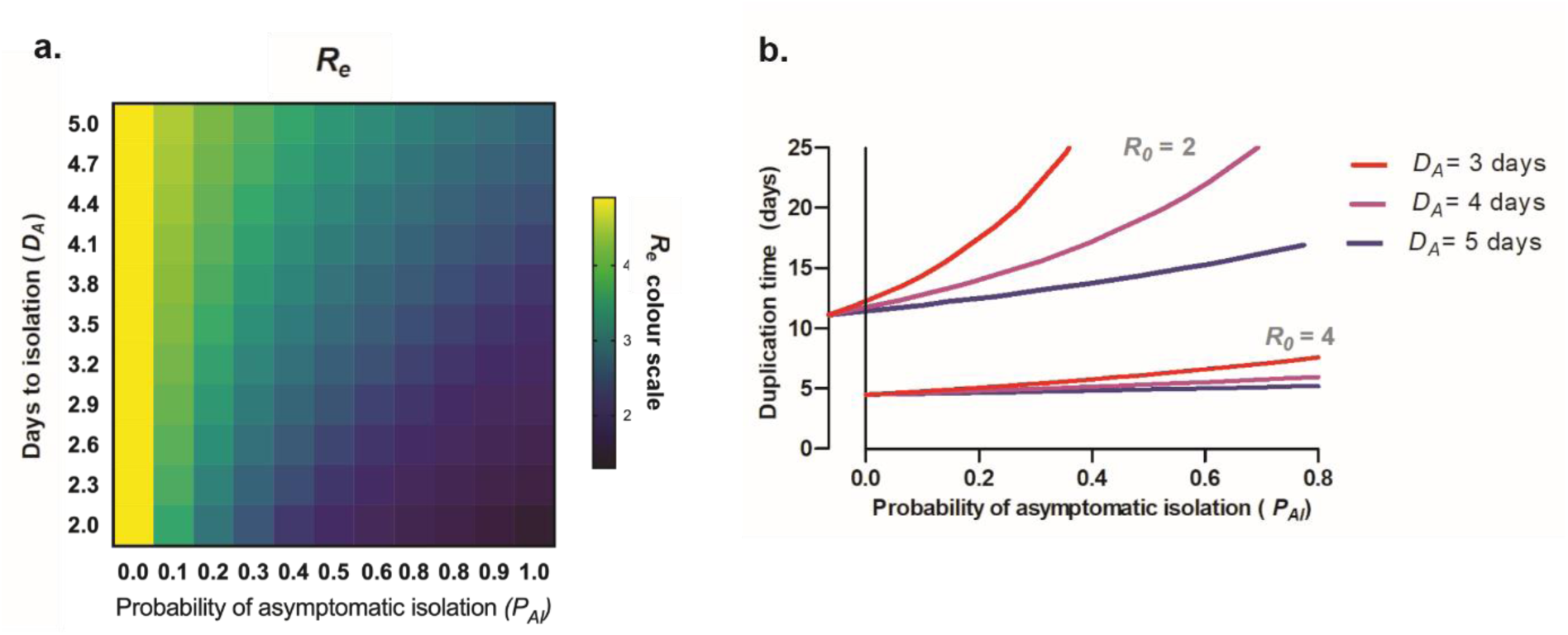
Effect of the probability of isolation of asymptomatic infectious individuals *(P_AI_)* at different times (day of asymptomatic isolation: *D_A_)* on the effective reproduction number *(R*_e_) (5a) and on the duplication time of the number of cases during the exponential phase of the epidemic (5b).

The consequence on the effective reproduction number of the removal of asymptomatic individuals from the infectious pool affects both epidemic dynamics and equilibrium values. The result is robust over a wide range of parameters. Fig. 6 shows the effect of efficient asymptomatic isolation on health capacity burden and overall mortality for the whole population. The plots show medians and interquartile ranges over a sample of 10000 sets of parameters. This set was built by sampling residence times from truncated normal distributions between 0 and twice the average value shown in table 2 with a standard deviation of 50% the average value. We assumed the detection and isolation of 50% of asymptomatic individuals in day 3.

**Fig. 6:**
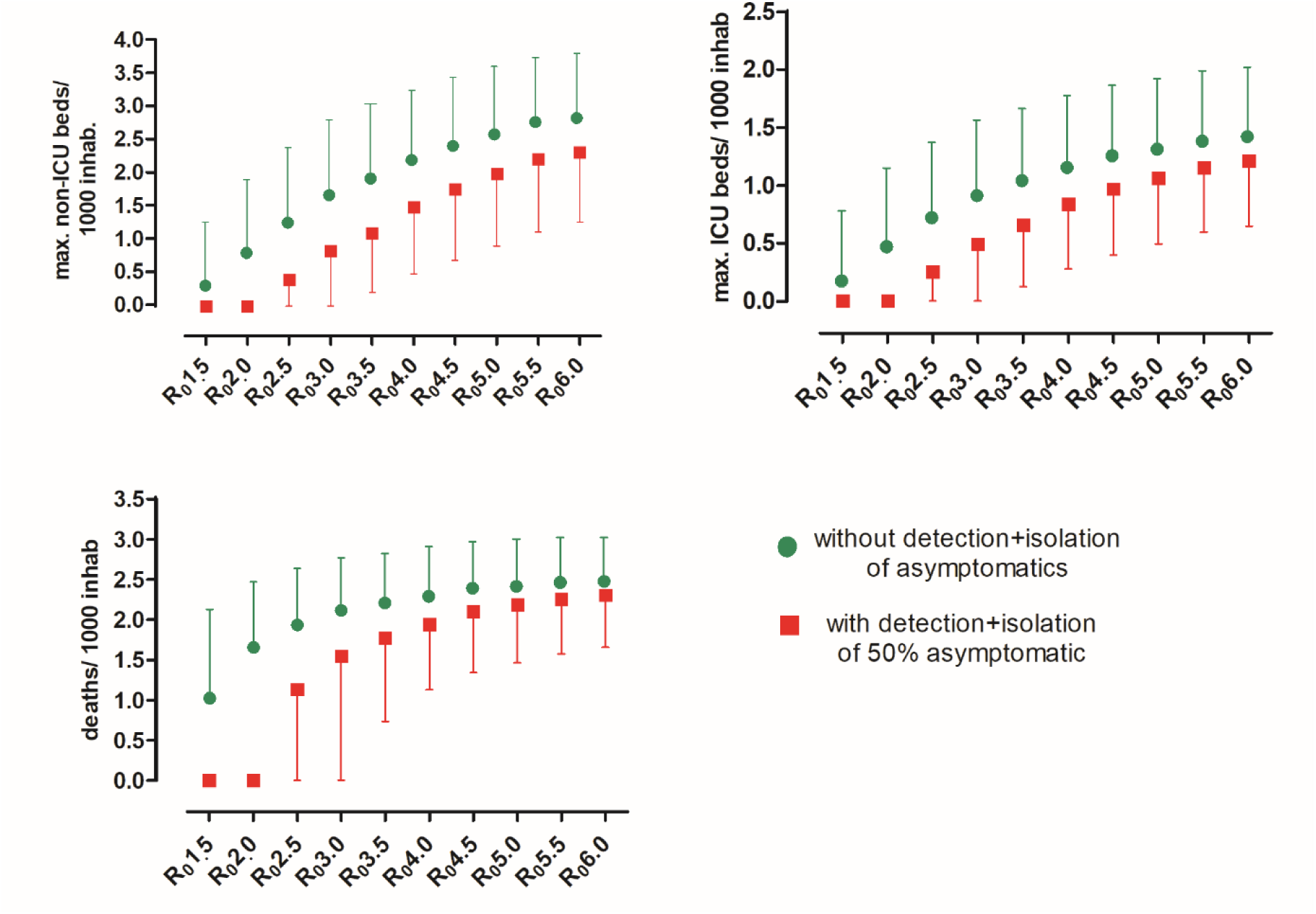
Healthcare burden effect of detection and isolation of 50% of asymptomatic individuals within three days of becoming infectious. The plots show medians and interquartile ranges over a sample of 10000 sets of parameters of important healthcare variables: maximum usage of hospital beds (ICU and non-ICU), and accumulated fatalities when system reaches equilibrium.

Efficient removal of asymptomatic infectious individuals from circulation has dramatic effects on healthcare burden and fatality over the total population, mainly when *R*_0_ remains at the lower end. Once more, this supports the importance to maintain other mitigation actions in combination with asymptomatic detection and isolation.

### Suppression Triggers

Considering the hospital beds and ICU facilities for each zone, we ventured to see if a zone-specific on-off suppression measure policy was feasible, as suggested by Ferguson et al[9]. Considering the results shown in Fig. 4, the communication between all the districts was diminished to a tenth as well. We set a trigger of 50% ICU occupancy (of beds destined to be used for COVID-19, assuming ~70% of total ICU beds were going to be intended to the epidemic) to turn on suppression measures and a stopper of 30% of ICU occupancy to relax these actions. We show results for an *R*_0_ = 4 fluctuating with an *R_t_* = 1 and an *R*_0_ = 2 combined with an *R_t_* = 1. Considering *R*_0_ = 4 as normal activities before this pandemic and *R*_0_ = 2 with necessary containment measures but not complete lockdown. An *R_t_* = 1 was considered when complete lockdown or quarantine was ordered.

In the first scenario, the exponentiality of an *R*_0_ = 4 curve is impossible to stop, even when a complete lockdown is set, and inevitably the sanitary system collapses in all areas, making this undoubtedly a lousy strategy. This evidence supports once more what we said in the previous scheme; life cannot return to normal when suppression measures are relaxed. (Fig. 7).

**Fig. 7:**
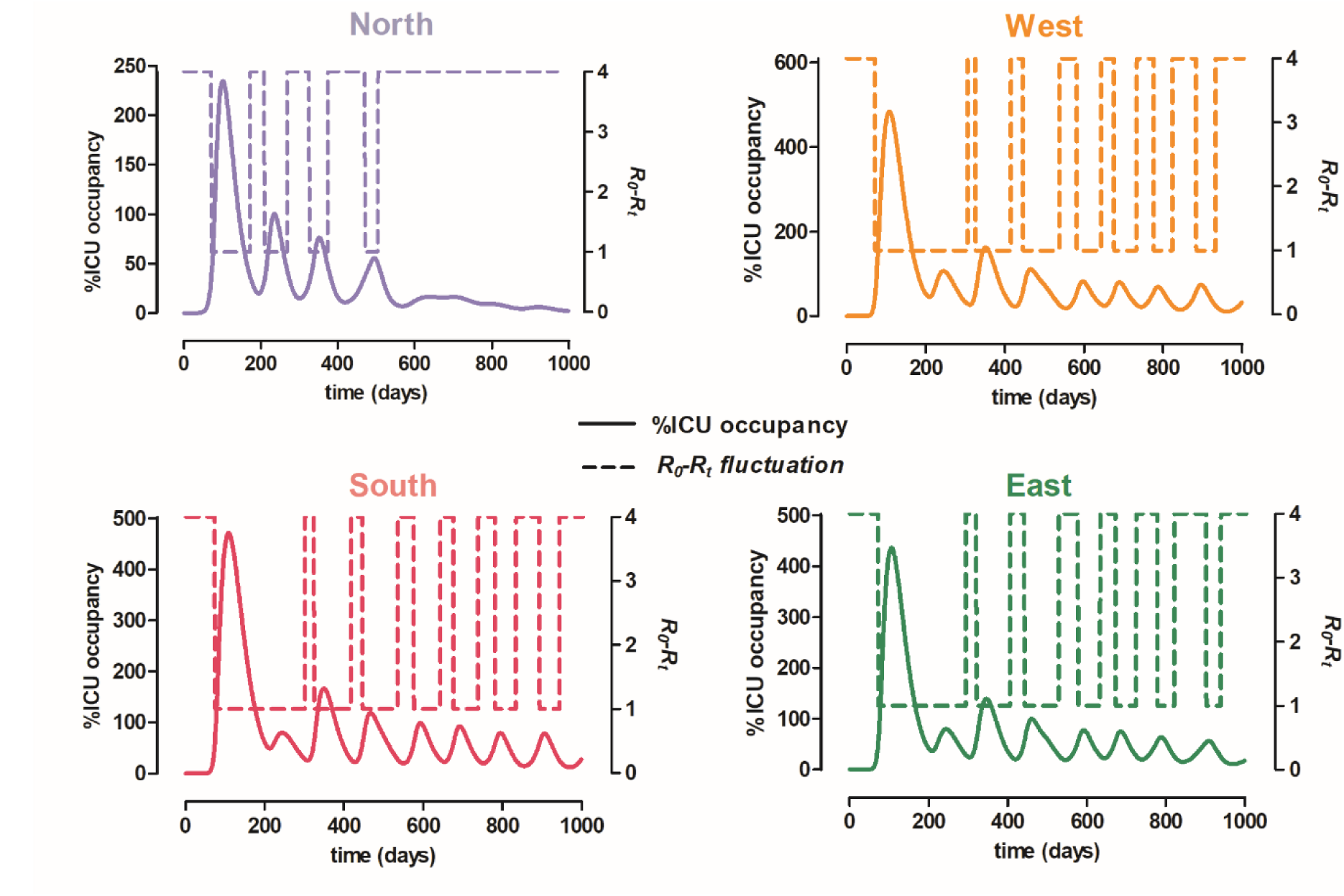
The effect of triggered on-off suppression measures in each region (trigger=50% ICU occupancy, stopper=30% ICU occupancy) in a *R*_0_=4 scenario. Dotted lines represent reproduction number fluctuance between *R*_0_=4 (no containment measures installed) and *Rt*=1 (lockdown). Full lines show % of ICU occupied beds throughout the timeline of 1000 days for which these on-off measures were simulated.

Considering that once the epidemic has struck, social standards are entirely changed, we simulated an alternation of *R*_0_ and *R*_t_ between 2 and 1, meaning this that once strict measures are relaxed, there are still essential actions being taken (i.e. social distancing, universities remain closed, etc.). In Fig. 8 we show that the Northern area can stand this situation without a sanitary collapse, needing two quarantines to surpass the epidemic. The other, more rural areas still have an overwhelmed health care system and need much more time in maximum isolation.

**Fig. 8:**
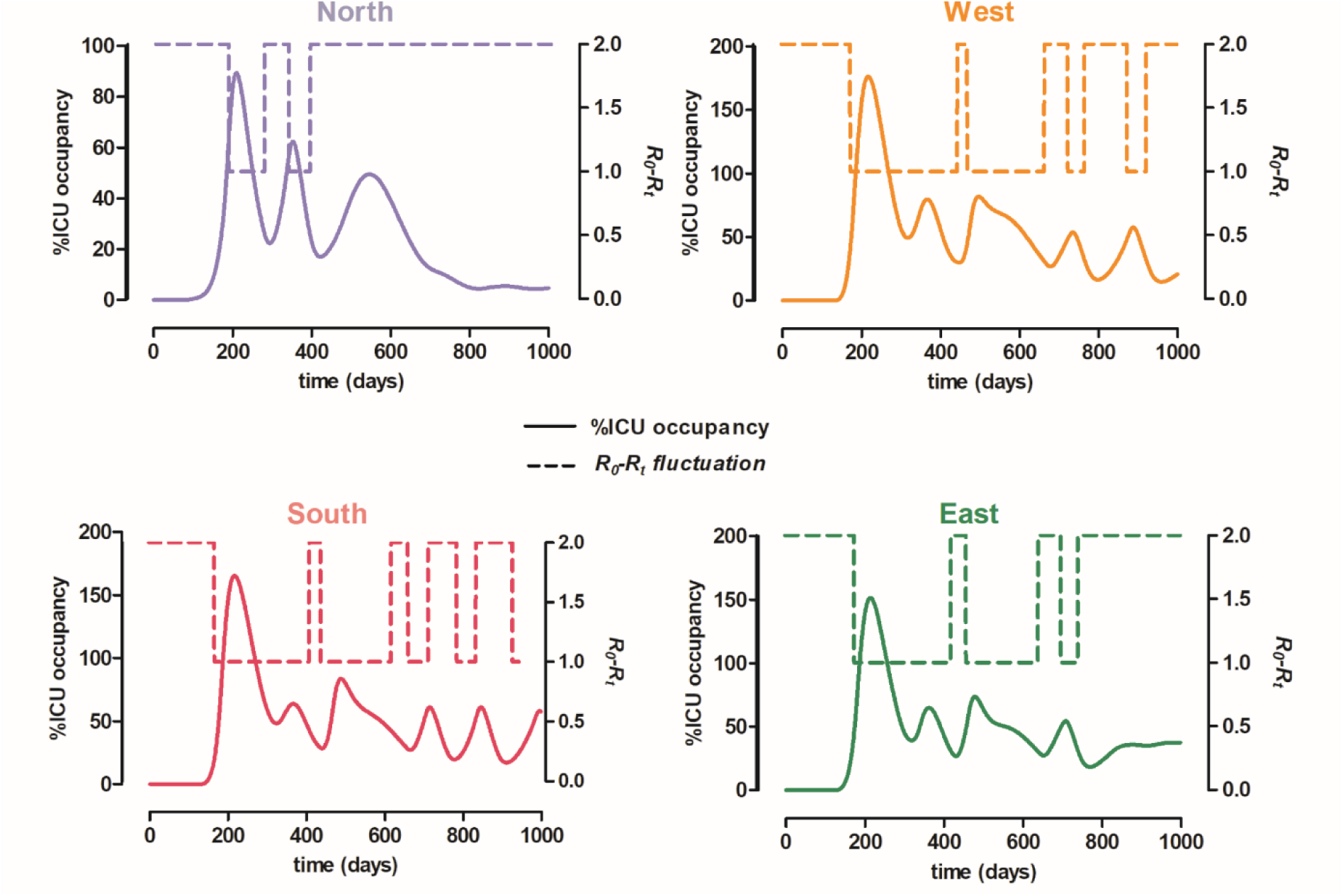
The effect of triggered on-off suppression measures in each region (trigger=50% ICU occupancy, stopper=30% ICU occupancy) in a *R*_0_*=2* scenario. Dotted lines represent reproduction number fluctuance between *R*_0_*=2* (moderate containment measures installed) and Rf1 (lockdown). Full lines represent % of ICU occupied beds throughout the timeline for which these on-off measures were simulated.

Since intensive health care facilities are mainly located in the North, we set our model with the ICU beds as a single pool for the whole province. We still maintained the areas separated regarding the rest of the parameters. Also, movement between cities remained restricted to a tenth. Contemplating this, we ran once more the model between *R_0_* = 2 and an *R_t_ =* 1. This strategy kept the health system below saturation and the quarantine periods were more acceptable for the whole territory. Still, in this scenario, 4.5 months of lockdown are needed to endure the pandemic and the inconvenience of having to transfer patients throughout the territory yet needs to be considered. (Fig. 9)

**Fig. 9:**
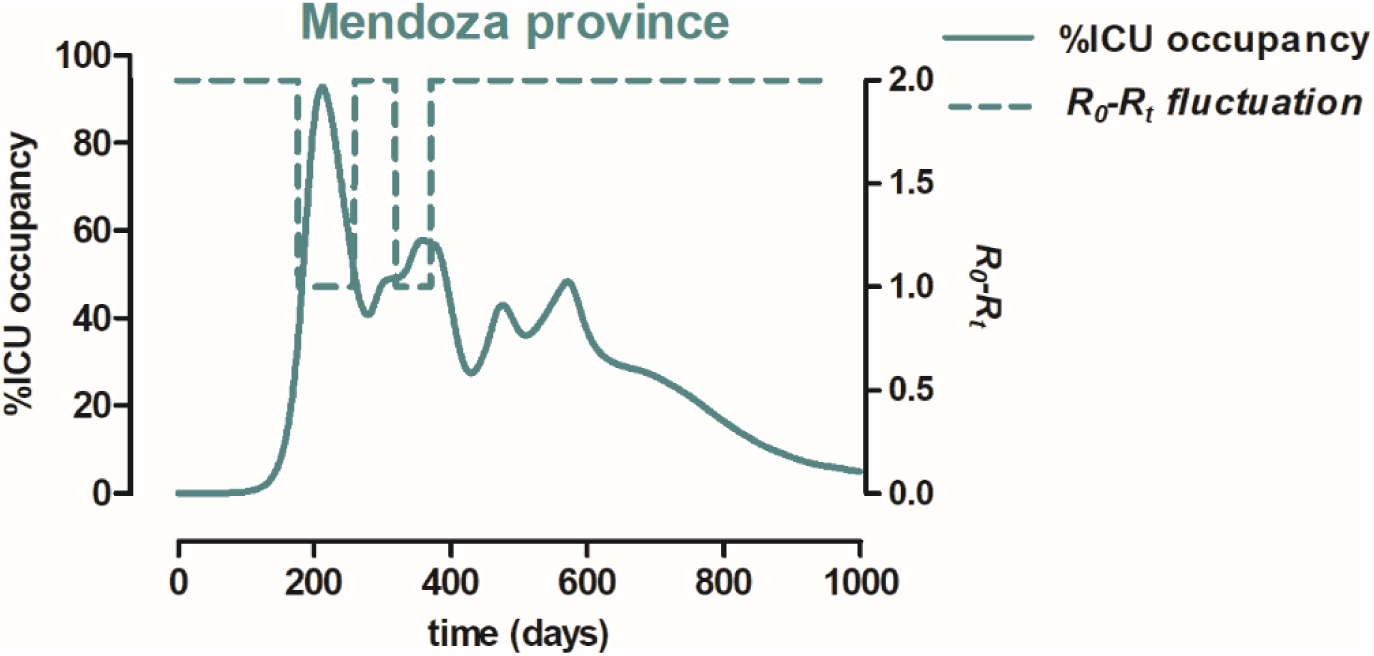
The effect of triggered on-off suppression measures in the whole territory (trigger=50% ICU occupancy, stopper=30% ICU occupancy) in a *R_0_* = 2 scenario. ICU beds were considered as a single pool for the entire province. Dotted lines represent reproduction number fluctuance between *R_0_* = 2 (moderate containment measures installed) and *R_t_* = 1 (lockdown). Full lines represent % of ICU occupied beds throughout the timeline for which these on-off measures were simulated.

Since the asymptomatic group is such an essential part of the system, we ventured to see what would happen if one could detect and isolate at least a proportion of them. Thus, using the same triggers as before (ICU beds 50%-30%), in a synchronized system, we added the detection and isolation of different proportions of asymptomatic individuals. In Fig. 10 we see that screening for asymptomatic cases diminishes significantly the time needed with complete lockdown. In the ideal assumption that 45% of the asymptomatics could be detected and isolated, there would be no need for quarantine, and the health care system would not collapse.

**Fig. 10:**
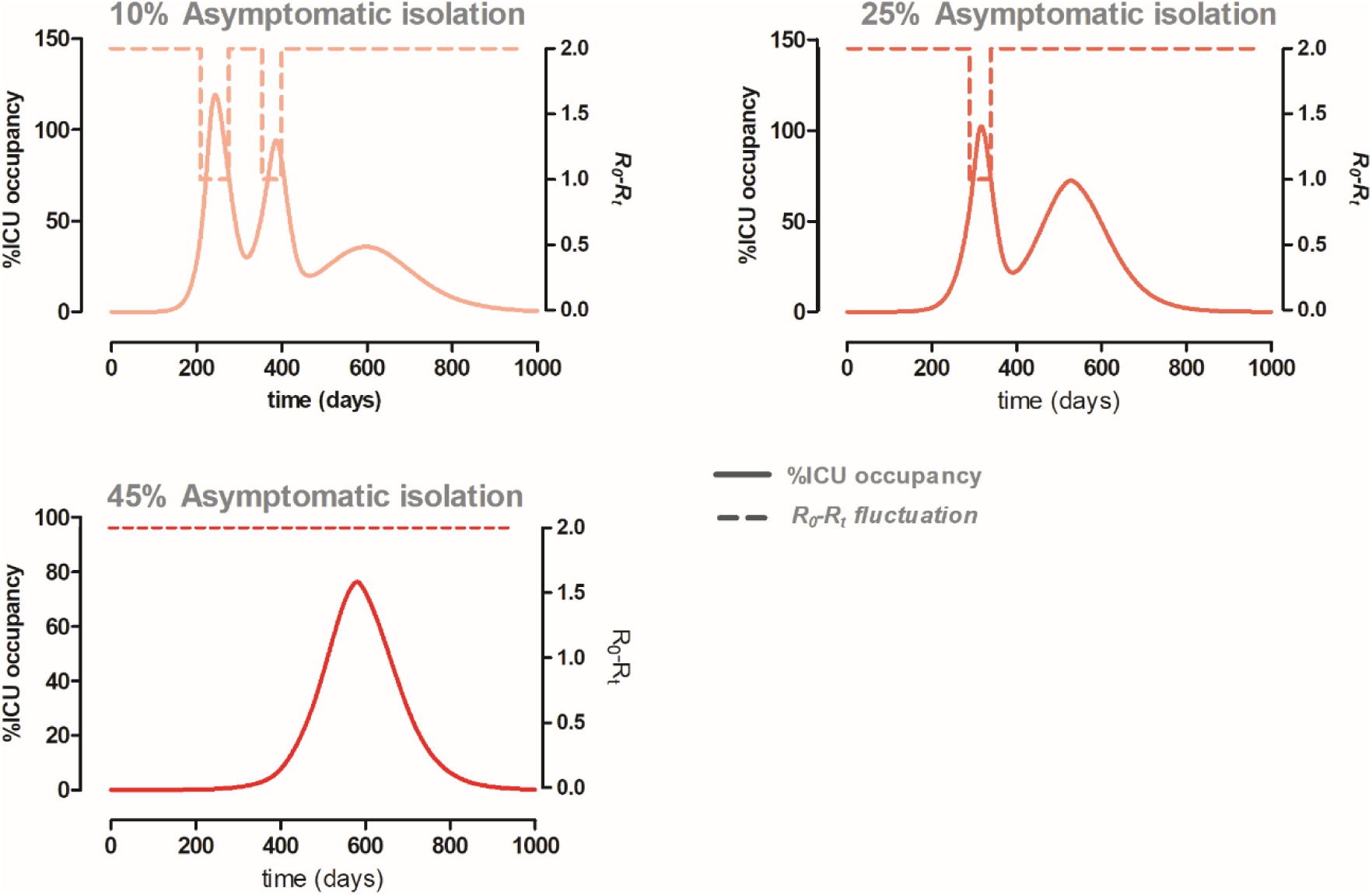
The effect of detecting and isolating different percentages of asymptomatic individuals on a model triggering on-off suppression measures in the whole territory (trigger=50% ICU occupancy, stopper=30% ICU occupancy) in a *R_0_* = 2 scenario. ICU beds were considered as a single pool for the entire province. Dotted lines represent reproduction number fluctuance between *R_0_* = 2 (moderate containment measures installed) and *R_t_* = 1 (lockdown). Full lines represent % of ICU occupied beds throughout the timeline for which these on-off measures were simulated.

## DISCUSSION AND CONCLUSIONS

Argentina was one of the countries that acted fastest and most rigorously very early in the arrival of the pandemic, setting a complete lockdown when only 128 cases had been reported. This policy has indeed "flattened the curve” as it has in many countries around the world for which the growth of clinical cases has changed from exponential to linear in time. The sustainment of drastic suppression measures over months, albeit possibly successful, is not sustainable and alternatives must be considered.

To establish a reliable mathematical model for a practically unknown disease was a challenge, but we had the advantage to count with previous epidemiological studies [3,11,13,14] and modelling proposals [7–9]. We established a locally inspired mathematical model of the disease, in the ~1,900,000 population province of Mendoza-Argentina, which can apply to any other city or country.

The model we propose provides explicit variables such as the number of patients under intensive care, hospital admissions, mild cases, and asymptomatic individuals. These variables are described dynamically according to the different residence times in each compartment and branching probabilities. We consider this to be a superior alternative to evaluating health burden parameters from a purely probabilistic point of view that does not consider the dynamical nature of the epidemic evolution. Furthermore, our model is adapted to the regional realities of small districts, which can be used as a strategy tool for any place in the world.

We confirmed that if an outbreak were to burst in one city, blocking circulating routes can as expected, have an essential impact. On the other hand, the health care distribution of Mendoza made a regionalized trigger for suppression impractical because of the uneven assignment of intensive care units. Still, in other countries or regions, this strategy might be useful.

The significant contribution we can make is to suggest that asymptomatic or very mildly ill patients provide a primordial hinge to manage the epidemic. Any control upon them exerts a substantial impact on the disease outcome. Previously described on-off suppression strategies [9] become much more effective when combined with the detection and isolation of asymptomatic cases. The association of mitigation measures with detection and isolation of around half of the asymptomatic and paucisymptomatic individuals would not need strict suppressive actions. Therefore, massive COVID-19 screening would be an alternative to the province’s complete lockdown.

For low-income countries, like ours, screening by pool-testing could be a helpful strategy if started early in the epidemic. Yelin et al. [15] showed that samples could be examined adequately in pools of 32, reducing dramatically the costs needed for extensive screening. Argentina could use this strategy to detect and isolate as many asymptomatic or very mildly affected individuals as possible to be able to reduce the time span over which strict suppression measures are in effect.

Our model and its analysis inform that the detection and isolation of *all* infected individuals, without leaving aside the asymptomatic group is the key to surpass this pandemic.

## Data Availability

A Python Jupyter Notebook implementing the model is available at a GitHub repository.

https://github.com/ihem-institute/SEIR_Mendoza

## Acknowledgements

The authors thank our affiliation institutions for supporting us and Mendoza’s Ministry of Health for encouraging and using this work.

## Data sharing

Code used in this study is available in the GitHub repository https://github.com/ihem-institute/SEIR_Mendoza

## Author contributions

CGS conducted the research project with the collaboration of LSM. CGS was in charge of the model design and the implementation of it with the help of LSM and GF. LM chose the epidemiological compartments and variables with the help of CGSam, SM and MVS. LM, CGS and LSM carried out simulations for the Mendoza province. LM and CGS wrote the manuscript and elaborated the figures. All authors have reviewed and approved the manuscript.

